# Quantification of the South African Lockdown Regimes, for the SARS-CoV-2 Pandemic, and the Levels of Immunity They Require to Work

**DOI:** 10.1101/2020.07.11.20151555

**Authors:** S. J. Childs

## Abstract

This research quantifies the various South African lockdown regimes, for the SARS-CoV-2 pandemic, in terms of the basic reproduction number, r_0_. It further calculates the levels of immunity required for these selfsame lockdown regimes to begin to work, then predicts their perceived values, should infections have been underestimated by a factor of 10. The first, level-5 lockdown was a valiant attempt to contain the highly infectious, SARS-CoV-2 virus, based on a limited knowledge. Its basic reproduction number (r_0_ = 1.93) never came anywhere close to the requirement of being less than unity. Obviously, it could be anticipated that the same would apply for subsequent, lower levels of lockdown. The basic reproduction number for the level-3 lockdown was found to be 2.34 and that of the level-4 lockdown, 1.69. The suggestion is therefore that the level-4 lockdown might have been marginally ‘smarter’ than the ‘harder’, level-5 lockdown, although its basic reproduction number may merely reflect an adjustment by the public to the new normal, or the ever-present error associated with data sets, in general. The pandemic’s basic reproduction number was calculated to be 3.16, in the Swedish context. The lockdowns therefore served to ensure that the medical system was not overwhelmed, bought it valuable time to prepare and provided useful data. The lockdowns nonetheless failed significantly in meeting any objective to curtail the pandemic.

## 1 Introduction

South Africa can be thought of as lucky, in that it obtained reasonably advanced warning of the impending pandemic, however, it is challenged in the way of its living conditions and basic hygiene. Many of its citizens live in extremely close proximity to one another and they are in constant physical contact.

The official narrative of the South African, SARS-CoV-2 epidemic is that it commenced around the 1st of March 2020, following the return of a group of seven, infected tourists, from a skiing holiday in Italy [6]. Multiple sources (e.g. [5] and [8]) subsequently reported that, by the 11th of March, the number of cases had risen to 13. By the 15th of March, there were already 61 reported cases ([10], [8] and [3]). By the 27th of March, a total of 1170 cases had been recorded and 1138 of them were still active ([10], [8] and [3]). A cursory inspection of this early data suggests that the first infection, in actual fact, took place substantially before March, or at very least, recoveries in early March were not properly reported.

The level-5 lockdown commenced on the 26th of March and ended on the 1st of May. Citizens were prohibited from leaving their residences for any other reason than to purchase food, electricity, fuel, or other items deemed essential. The rules only allowed foodstuffs and other essential goods to be sold. Limits on the numbers of customers, in the same shop at the same time, were also set. Only those employed in supermarkets, medical personnel, emergency plumbers and their like were allowed to continue working. The sale of tobacco and liquor was also prohibited. Further details of the level-5 lockdown regulations are available at [7]. The opening weeks of the level-5 lockdown were characterised by migration and a substantial lack of compliance. This is borne out by, for example, a plethora of photographs showing traffic jams at Hertzog Bridge, in Aliwal North, and videos showing kilometres-long convoys of minibuses, at other Eastern Cape borders; all of which appeared on social media. The flagrant disregard for the rules was not limited to migrant workers and holiday makers. The elite of at least one establshed institution had guests around, tasted and distributed homemade liquor among themselves, had workmen in and sent their children to each others’ houses. In many areas, the large crowds that queued outside supermarkets suggested that people needed time to adjust to the level-5 rules. People in many of these queues were in physical contact, or very close to it. By the 1st of May, the day after the level-5 lockdown had ended and the day the level-4 lockdown commenced, a total of 5951 cases had been recorded and 3453 of those were still active ([10], [8] and [3]).

The level-4 lockdown commenced on the 1st of May and ended on the 31st day of that same month. The level-4 rules allowed for exercise in public places, between the hours of 06h00 and 09h00, stipulated that people must wear masks, at all times, in public and a travel ban was implemented on the 7th of May. Further details of the level-4 lockdown regulations are available at [7]. This lockdown was characterised by fewer infringements, as the public appeared to become adjusted to the new normal. By the 31st of May, at the end of the level-4 lockdown, a total of 32683 cases had been recorded and 15191 of those were still active ([10], [8] and [3]).

The level-3 lockdown commenced on the 1st of June and was still in force on the first day of that following month. The level-3 rules permitted the sale of liquor and, on the 8th of June, grades 7 and 12 returned to school. On the night of the 17th of June, so-called “advanced level-3” was announced. Hairdressers, restaurants, accredited and licenced accommodation, cinemas and casinos were notified that they could resume operating. How long they took to respond and to what level customers resumed patronising these establishments, is not known.

Further details of the level-3 lockdown regulations are available at [7]. By the 1st of July, a total of 159333 cases had been recorded and 80559 of those were still active ([10], [8] and [3]).

Sweden set no special regulations for their SARS-CoV-2 epidemic and their data ([11] and [9]) therefore serve as a convenient, nonetheless, very approximate control for South Africa’s lockdown experiments. Sweden’s climate, their population’s way of life, their population-densities etc. are, of course, all very different to those in South Africa, so one has to exercise caution in drawing any conclusions from the comparison.

The basic reproduction number, *r*_0_, may be defined as the total number of people that an infected member of the population would manage to infect, before recovering, in an otherwise naive population. Its significance is, however, far greater. In the basic reproduction number one has a holistic quantity with which to characterise both the infectiousness of a disease, as well as the environment in which it propagates, right down to things like the temperature of nasal passages, the rules of a lockdown and even the level of non-compliance. It allows for the lockdown-threshold to be calculated, which is the minimum level of immunity the population must have in order for the epidemic to downgrade to a mere disease, as well as the level at which all infection ceases. In one Chinese context, the basic reproduction number for the SARS-CoV-2 virus was found to be 2.2 [21].

In this research, the basic reproduction number, associated with each lockdown regime, is calculated from the numbers of active infections and the total tallys of all infections since the outbreak of the pandemic. Not only have so-called individual level models, such as those of [17] and [19], been discredited by [19] as a means for calculating *r*_0_, they are also far more laborious than the simple method used in this work. The various lockdown-thresholds, as well as the points at which all infection would cease, are, in turn, calculated from these basic reproduction numbers. Finally, the perceived values of these selfsame quantities are predicted, should infections have been underestimated by a factor of 10 and these results are compelling.

## 2 Derivation of the Relevant Formulae

Kermack and McKendrick’s SIR equations [20] state that

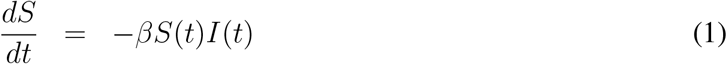

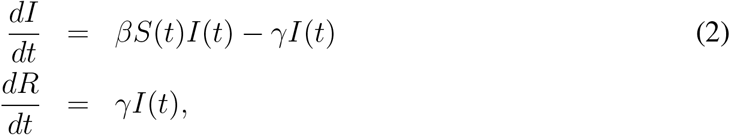

in which *β* denotes the rate of potentially infectious encounters to which a member of the population is exposed, *S*(*t*) denotes the susceptible fraction of the population, *I*(*t*) denotes the infected fraction of the population, *γ* denotes the combined rate of recovery and death, while *R*(*t*) denotes the ‘resistant’ fraction of the population, those that have either acquired immunity, or died from the disease. The characterisation of an epidemic in terms of a basic reproduction number, *r*_0_, the calculation of *r*_0_, the threshold and the point at which an infection completely burns itself out, are ultimately all based on these equations.

**Theorem 1 (Basic Reproduction Number)** *An epidemic is only possible for r*_0_ *>* 1.

**Proof:** By definition, a disease is not an epidemic unless the level of infection increases. From Equation (2), one observes

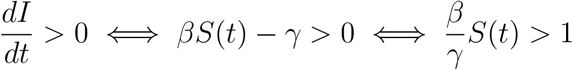

Since *S*(*t*) *≤* 1 for all *t*,

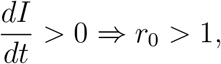

in which 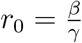, concluding the proof.

In *r*_0_ one therefore has a holistic quantity with which to characterise both the infectiousness of a disease, as well as the environment in which it propagates, right down to things like the temperature of nasal passages, the rules of a lockdown and even the level of non-compliance. The so-called *r*-effective, *r*_0_ *× S*(*t*), is a characterisation of the disease’s infectiousness, pertinent to a given point in time, as the epidemic progresses, or where immunity is present.

### A Formula With Which to Recover the Basic Reproduction Number From the Data

Using the chain rule,

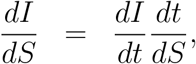

then substituting Equations (2) and (1), the expression,

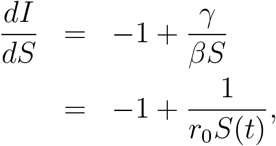

is obtained. Integrating over S,

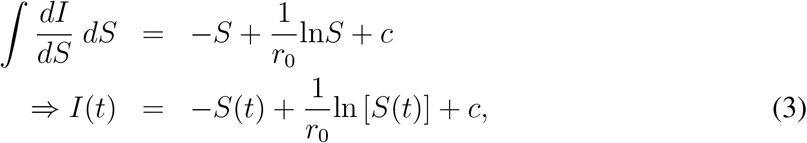

is obtained, in which *c* is the unknown constant of integration. At some *t* = *t*_*i*_,

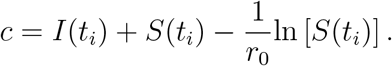

Quantification of Lockdown Regimes

Substituting this back into Equation (3),

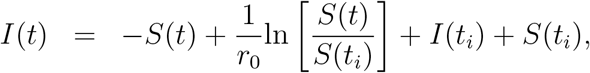

is obtained. Evaluating this equation over some time interval [*t*_1_, *t*_2_] and solving for *r*_0_,

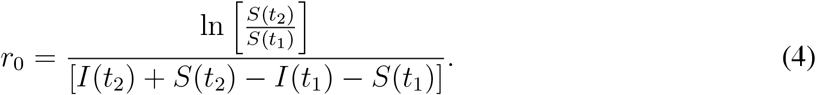

Notice that this formula is robust against any movement of *I*(*t*) by an additive constant, up or down. Such movement has no effect on the calculation of *r*_0_, is reasonably robust against any data error that does not effect the relative values, or slopes of the functions concerned.

Once *r*_0_ has been calculated, *S*(*t*_2_) = *S*_*∞*_, can be recovered from this selfsame equation, by considering that *S*_*∞*_ is the point at which all infection ceases, i.e. by setting *I*(*t*_2_) = 0, in the above equation. Once the level of infectiousness for a given lockdown has been characterised in terms of the basic reproduction number, *r*_0_, it is instructive to know the level of immunity at which the lockdown renders the disease no longer an epidemic. The levels to which susceptability must drop, in order for the relevant lockdown regimes to begin to work, can be determined by the application of the threshold theorem.

**Theorem 2 (The Threshold Theorem)** *No epidemic occurs when* 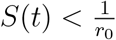.

**Proof:** By definition, a disease is not an epidemic if the level of infection decreases. From Equation (2), one observes

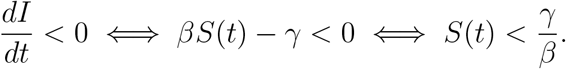

Since the quantity on the right is immediately recogniseable as 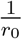, the above statement may be more concisely expressed as,

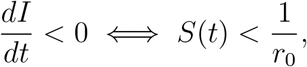

concluding the proof.

In other words, if the susceptable fraction of the population is still above 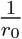, for a given lockdown, that lockdown will not serve to curtail the epidemic, only to delay it. In such circumstances, the epidemic will only temporarily slow during the lockdown, then resume, as before, after it. Only at the threshold does *r*-effective drop to unity.

## 3 Asymptomatic, or Undiagnosed SARS-CoV-2 Infections

What of the large number of asymptomatic, or undiagnosed SARS-CoV-2 infections, not reflected in the data? The head of the CDC, Robert Redfield’s opinion on the topic of asymptomatic or undiagnosed SARS-CoV-2 infections, in the U.S.A., is that antibody testing reveals that “A good rough estimate now is 10 to 1” [4] and others, in similar positions all over the world, have expressed similar sentiments.

Suppose one were to determine that both *I*(*t*) and therefore, the resistant portion arising from the current epidemic, *R*(*t*) *− R*(0), are in actual fact higher by a factor, *a*. That is,

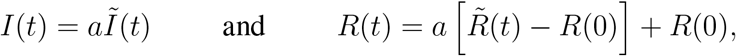

in which the tilde denotes the incorrectly measured, data-value in an epidemic that begins at *t* = 0. Then

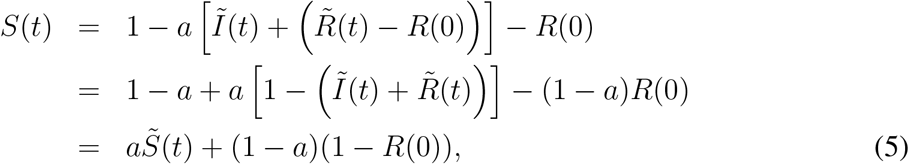

in which 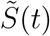 denotes the perceived, incorrect-data-based value of *S*(*t*). The threshold therefore occurs at

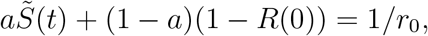

according to Theorem 2, from which the corresponding, perceived, incorrect-data-based threshold can be recovered by changing the subject of the above equation to

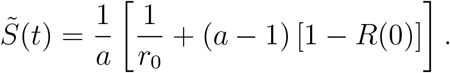

Changing the subject of Equation (5) also facilitates the calculation of the perceived, incorrect-data-equivalent of *S*_*∞*_ (*t*),

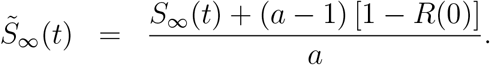

Of course, *R*(0) = 0 for a novel infection such as SARS-CoV-2; or so one believes.

## 4 Fitting Curves to the Data

Curves are fitted to a level-5 data set, a level-4 data set and a Swedish data set. Epidemiological data are usually presented in the format “numbers of current infections” and “total number of cases”. The present case of the SARS-CoV-2 pandemic is no exception. If *N* is the size of the population, the aforementioned quantities are just *I*(*t*)*N* and [*I*(*t*) + *R*(*t*)]*N*, respectively; always assuming that the population is naive in the case of a novel infection like SARS-CoV-2 (i.e. that *R*(0) = 0). This standard, epidemiological data format merely implies an additional step; namely that the values S(t) and I(t), to be used in the formulation (4), must first be calculated from

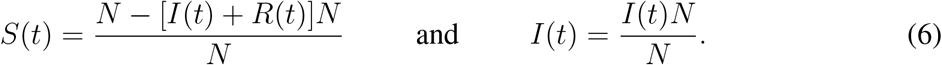

In 2020, population sizes were estimated to be 59 140 502 and 10 089 108 for South Africa [12] and Sweden [13], respectively.

### The Applicable Temporal Interval

The mean incubation period for the SARS-CoV-2 virus is 5.2 days with the 95th percentile occurring at 12.5 days [21]. The World Health Organisation (WHO) quotes the incubation period as being anywhere between 2 and 10 days [15], China’s National Health Commission (NHC) found symptoms to appear anywhere from 10 to 14 days after infection [1], the United States’ Centres for Disease Control and Prevention (CDC) found symptoms to appear anywhere from 2 to 14 days after infection [16] and the Chinese, online DXY.cn quotes the incubation period as being anywhere between 3 to 7 days after infection; possibly as high as 14 days [14]. Two, record outliers for the incubation period are 19 days [18] and 27 days [2]. There is therefore considerble agreement on a lower bound of no less than 2 days and an upper bound of no more than 14 days for the incubation period. Allowing a further day for diagnosis, data were used from the sixteenthth day after the relevant lockdown began until the first day after it ended.

#### 4.1 The Level-5 Lockdown

The level-5 lockdown commenced on the 26th of March and ended on the 1st of May. A period of 15 days was allowed for the viral incubation period and the subsequent diagnosis of an infection. It was also assumed that the termination of the level-5 lockdown would not reflect in the data for at least 24 hours.

Curves were accordingly fitted to the subset of data ([10], [8] and [3]) which commenced on the 10th of April and terminated on the 1st of May. The curves fitted to the data, using Gnuplot, are depicted in Figure 1. The formula for the “total infections” curve is

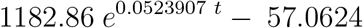

and the formula for the “active infections” curve is

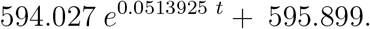

**Figure 1:**
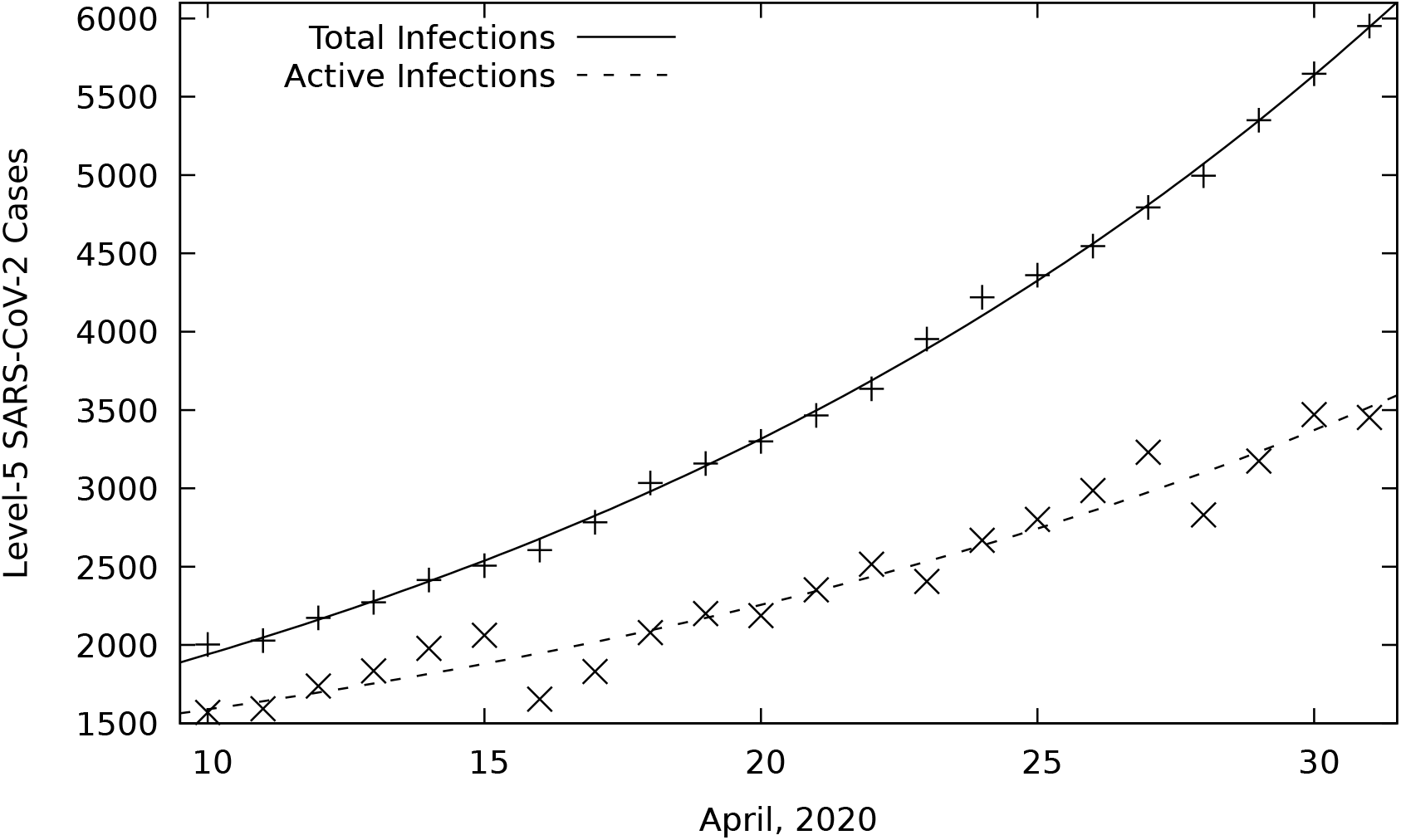
Level-5, best fits to SARS-CoV-2, infection data (10th of March to the 1st of May, 2020).

The values these formulae yield are provided in Table 1. They are first substituted into Equations (6), which, in turn, provide the necessary inputs for Equation (4).

**Table 1:**
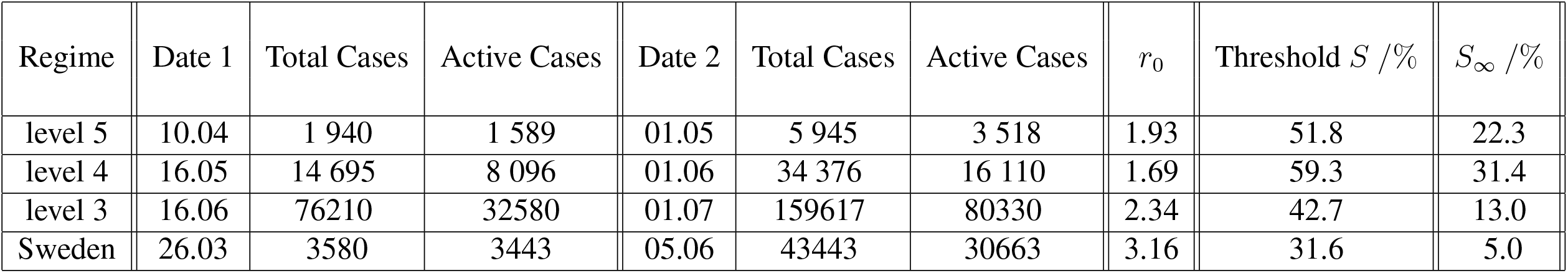
The basic reproduction number, r_0_, the consequent threshold and the point at which all infection ceases, S_∞_. With the exception of Sweden, all are calculated from data associated with the relevant lockdown regimes, imposed for the SARS-CoV-2 pandemic, in South Africa.

| Regime | Date 1 | Total Cases | Active Cases | Date 2 | Total Cases | Active Cases | $r_0$ | Threshold $S$ /% | $S_\infty$ /% |
| --- | --- | --- | --- | --- | --- | --- | --- | --- | --- |
| level 5 | 10.04 | 1 940 | 1 589 | 01.05 | 5 945 | 3 518 | 1.93 | 51.8 | 22.3 |
| level 4 | 16.05 | 14 695 | 8 096 | 01.06 | 34 376 | 16 110 | 1.69 | 59.3 | 31.4 |
| level 3 | 16.06 | 76210 | 32580 | 01.07 | 159617 | 80330 | 2.34 | 42.7 | 13.0 |
| Sweden | 26.03 | 3580 | 3443 | 05.06 | 43443 | 30663 | 3.16 | 31.6 | 5.0 |

#### 4.2 The Level-4 Lockdown

The level-4 lockdown commenced on the 1st of May and ended on the 31st of May. Once again, a period of 15 days was allowed for the viral incubation period and the subsequent diagnosis of an infection. Once again, it was also assumed that the termination of the level-4 lockdown would not reflect in the data for at least 24 hours. Curves were accordingly fitted to the subset of data ([10], [8] and [3]) which commenced on the 16 of May and terminated on the 1st of June. The curves fitted to the data, using Gnuplot, are depicted in Figure 2.

**Figure 2:**
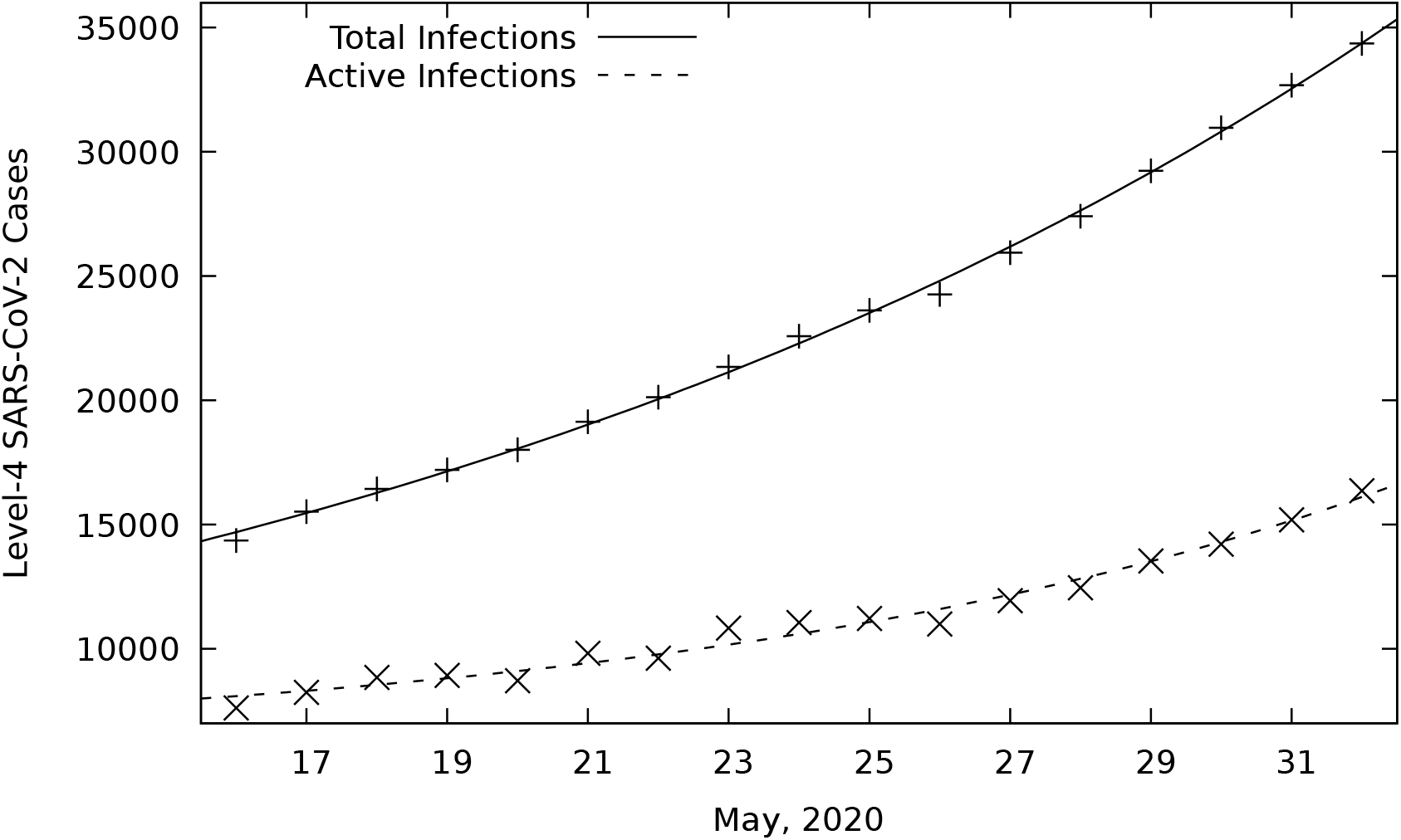
Level-4, best fits to SARS-CoV-2, infection data (16th of May to the 1st of June, 2020).

The formula for the “total infections” curve is

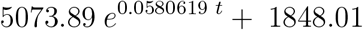

and the formula for the “active infections” curve is

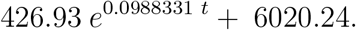

The values these formulae yield are provided in Table 1. They are first substituted into Equations (6), which, in turn, provide the necessary inputs for Equation (4).

#### 4.3 The Level-3 Lockdown

The level-3 lockdown commenced on the 1st of June and was still in force one month later. Once again, a period of 15 days was allowed for the viral incubation period and the subsequent diagnosis of an infection. Curves were accordingly fitted to the subset of data ([10], [8] and [3]) which commenced on the 16 of June and terminated on the 1st of July. The curves fitted to the data, using Gnuplot, are depicted in Figure 3.

**Figure 3:**
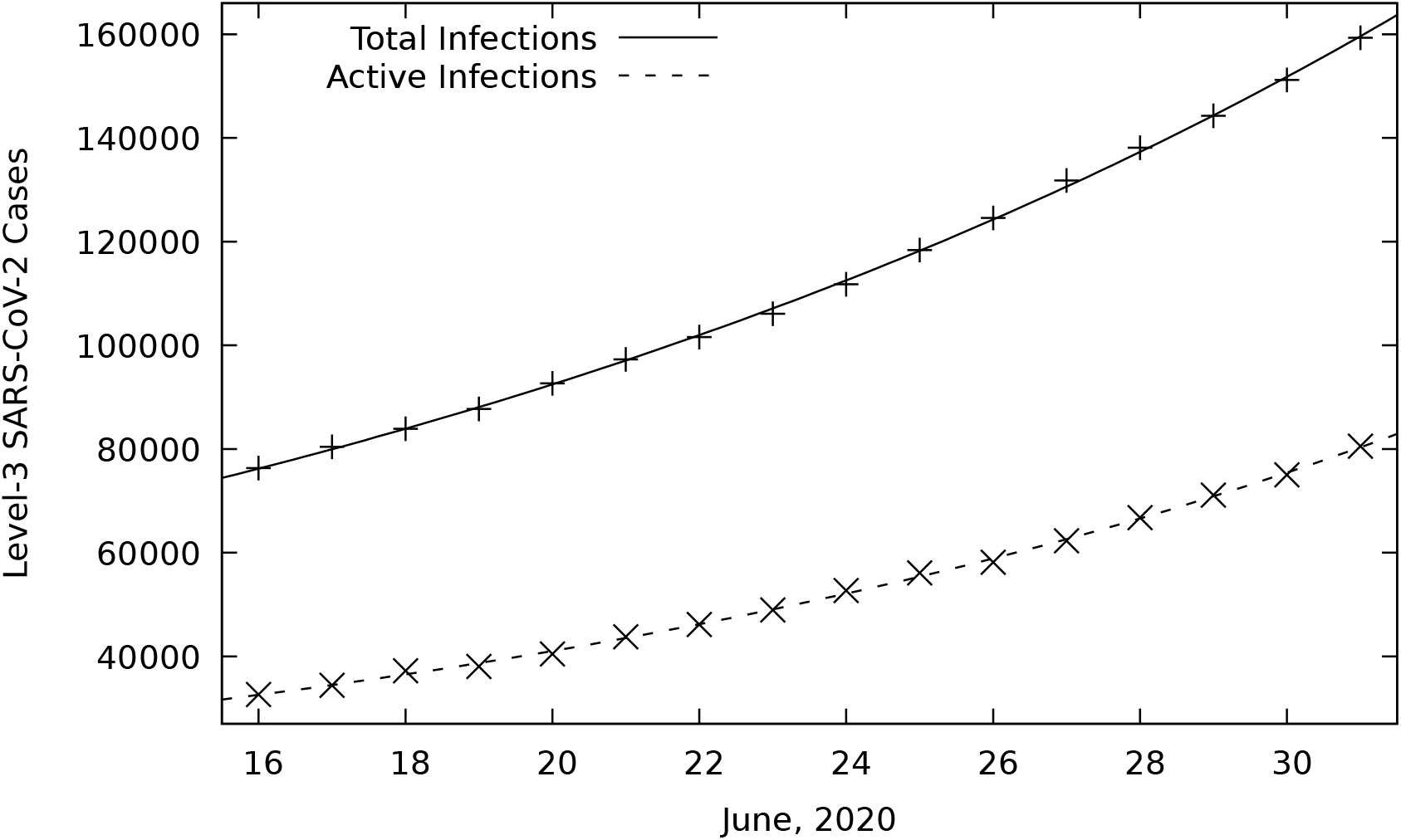
Level-3, best fits to SARS-CoV-2, infection data (16th of June to the 1st of July, 2020).

The formula for the “total infections” curve is

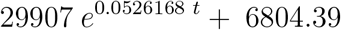

and the formula for the “active infections” curve is

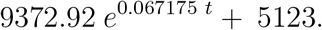

The values these formulae yield are provided in Table 1. They are first substituted into Equations (6), which, in turn, provide the necessary inputs for Equation (4).

#### 4.4 Sweden

Sweden’s data [11] appear to be regularly subjected to major revisions. The data used in this work might therefore no longer be current at the time of reading, however, the actual values of the data and consequentlly the results, change little. The regular revisions suggest that someone cares.

Visual inspection of the Swedish data revealed a much less prominent, initial step, up. It is fairly safe to assume this would be indicative of discovery, rather than growth of the actual SARS-CoV-2 pandemic, itself, as they had no lockdowns. Sweden’s data ([11]) are slightly problemmatic, in that an uncharacteristically large number of recoveries were reported on the 4th of May and the 2nd of June. The synchronicity in these recoveries is highly suspect and they manifest themselves as two, visibly large steps, down, in the graph of “active infections” (Figure 4). One suspects that these ‘saw-teeth’ are, in actual fact, artefacts, which arose due to a delay in the diagnosis and reporting of recoveries, a suspicion which seemed to originally be corroborated by [11]. It is strongly suggestive of an incorrect record of “active infections” prior to the day in question. A decision was therefore made to fit curves to as much data as possible, barring early March (for fear that it documented discovery, rather than growth). Of course, one has to wonder whether the data subsequent to the 3rd of May and the 2nd of June were not also afflicted by a similar hoarding of recoveries and, indeed, they seem to be [11]. The ‘saw-teeth’ had the effect of ‘lifting’ the curve slightly, however, the overall slope appeared to be and, indeed, should mostly be correct for the temporal period selected. Fortunately, the formula for *r*_0_, Equation (4), is robust against any movement of *I*(*t*) by an additive constant, up or down. The *r*_0_ calculated according to the formula, Equation (4), does not change in any way, whatsoever, for such movement of *I*(*t*). A corrected curve, obtained by revising the original fit downward by 1100 infections, is nonetheless provided for the reader’s benefit, in Figure 4.

**Figure 4:**
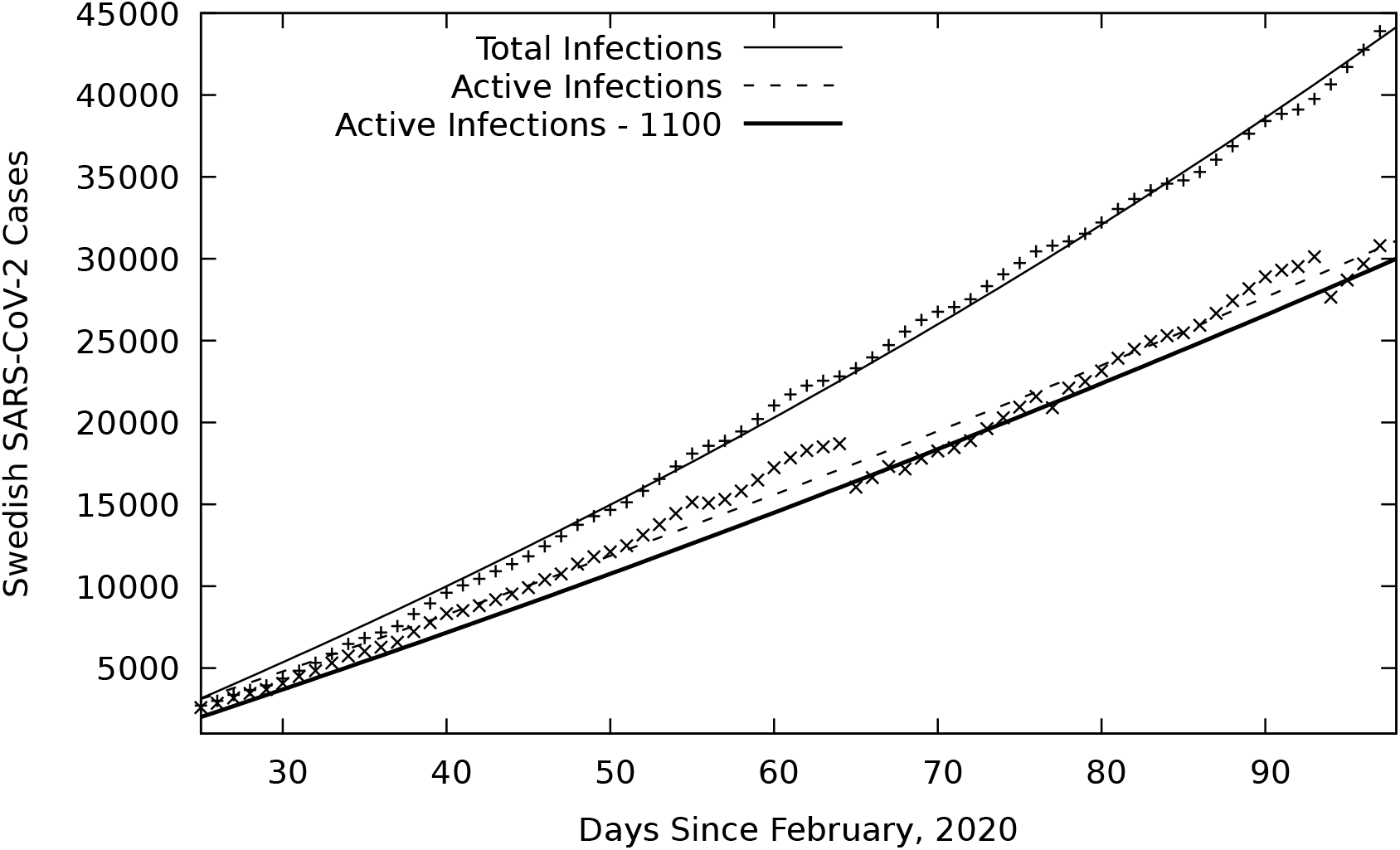
Best fits to the Swedish, SARS-CoV-2, infection data, from the 15th of March to the 5th of June, 2020.

The curves fitted to the subset of data ([11] and [9]), which commenced on the 15th of March and terminated on the 5th of June, are depicted in Figure 4. The curves were fitted using Gnuplot. The formula for the “total infections” curve is

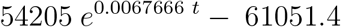

and the formula for the “active infections” curve is

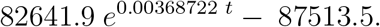

The values these formulae yield are provided in Table 1. They are first substituted into Equations (6), which, in turn, provide the necessary inputs for Equation (4).

## 5 Results

The results, as well as the inputs from which they were obtained, are provided in Table 1, on page 12.

## 6 Conclusions

There is very little difference between perceived and actual basic reproduction numbers, should infections have been underestimated by a factor as high as 10. They rise only very slightly. There is, however a profound difference between the actual and perceived, data-based thresholds and the susceptability levels at which the infection will vanish (comparing Tables 1 and 2). The results based on the assertion that infections have been underestimated by a factor of 10, are compelling.

**Table 2:** The basic reproduction number, r_0_ for a = 10, the consequent perceived threshold and the perceived point at which all infection ceases. With the exception of Sweden, all are calculated from data associated with the relevant lockdown regimes, imposed for the SARS-CoV-2 pandemic, in South Africa.

| Regime | Date 1 | Total Cases<sup>†</sup> | Active Cases<sup>†</sup> | Date 2 | Total Cases<sup>†</sup> | Active Cases<sup>†</sup> | $r_0$ | Threshold $\tilde{S}$ /% | $\tilde{S}_\infty$ /% |
| --- | --- | --- | --- | --- | --- | --- | --- | --- | --- |
| level 5 | 10.04 | 1 940 | 1 589 | 01.05 | 5 945 | 3 518 | 1.93 | 95.2 | 92.2 |
| level 4 | 16.05 | 14 695 | 8 096 | 01.06 | 34 376 | 16 110 | 1.69 | 95.9 | 93.1 |
| level 3 | 16.06 | 76 210 | 32 580 | 01.07 | 159 617 | 80 330 | 2.39 | 94.2 | 91.2 |
| Sweden | 26.03 | 3580 | 3443 | 05.06 | 43443 | 30663 | 3.23 | 93.1 | 90.5 |

<sup>†</sup> The perceived, incorrect data.

Although the initial attempt to contain the epidemic failed, the sequence of lockdowns provided vital data to determine their associated basic reproduction numbers and, consequently, at what levels of immunity these selfsame lockdowns would begin to become effective in the future (their associated epidemic-thresholds).

Early data, collected prior to the level-5 lockdown, probably document the trajectory of discovery, rather than they do growth of the actual SARS-CoV-2 pandemic, itself. This conclusion is based on a discernable step, up, in the graph, coupled with an observation that such phenomenal growth would have implied an unrealistic basic reproduction number; one of somewhere around 36! At very least, recoveries in the early data were under-reported. A lack of confidence in the very early data, along with abrupt transitions in lockdown regimes and their consequent inflection points (delayed, diffuse or otherwise), necessitated that curves only be fitted locally.

The first, level-5 lockdown was a valiant attempt to contain the highly infectious, SARS-CoV-2 virus, based on a limited knowledge. Its basic reproduction number (*r*_0_ = 1.93) never came anywhere close to the requirement of being less than unity. To put this in context, most influenza epidemics have a substantially lower *r*_0_ than this virus has under the conditions of a level-5 lockdown. Such a level-5 lockdown would only become efficacious in curtailing the SARS-CoV-2 pandemic, were it to be implemented around the 51.8 % susceptability level (Table 1). In other words, only after 48.2 % of the population has been infected. Under level-5 lockdown conditions, the SARS-CoV-2 virus would only vanish as a disease, forever, at around the 22.3 % susceptability level (Table 1). If, however, the assertion that infections have been underestimated by a factor of 10 is correct, then the susceptability level for the above threshold will be perceived as 95.2 %, while the susceptability level at which the infection will vanish, forever, will be perceived as 92.2 % (Table 2).

Obviously, it could be anticipated that the basic reproduction number for subsequent, lower levels of lockdown would also not come anywhere near the requirement of being less than unity. Indeed, the basic reproduction number for the level-3 lockdown was found to be 2.34 and that of the level-4 lockdown, 1.69 (Table 1). It is nonetheless surprising that the value characterising level-4 is not higher than that chracterising level-5. It could reflect an adjustment by the public to the ‘new normal’, the wearing of masks, the travel ban, or any combination of these. The suggestion is therefore that the level-4 lockdown was ‘smarter’ than the ‘harder’ level-5 lockdown, although the data may not be of sufficient quality to draw such a conclusion.

The level-4 lockdown would only be efficacious in curtailing the pandemic, were it to be implemented around the 59.3 % susceptability level (Table 1). In other words, only after 40.7 % of the population has been infected. Under level-4 lockdown conditions, the SARS-CoV-2 virus would only vanish as a disease, forever, at around the 31.4 % susceptability level (Table 1). If, however, the assertion that infections have been underestimated by a factor of 10 is correct, then the susceptability level for the above threshold will be perceived as 95.9 %, while the susceptability level at which the infection will vanish, forever, will be perceived as 93.1 % (Table 2).

The level-3 lockdown would only be efficacious in curtailing the pandemic, were it to be implemented around the 42.7 % susceptability level (Table 1). In other words, only after 57.3 % of the population has been infected. Under level-3 lockdown conditions, the SARS-CoV-2 virus would only vanish as a disease, forever, at around the 13.0 % susceptability level (Table 1).

If, however, the assertion that infections have been underestimated by a factor of 10 is correct, then the susceptability level for the above threshold will be perceived as 94.2 %, while the susceptability level at which the infection will vanish, forever, will be perceived as 91.0 % (Table 2).

In the Swedish context, the basic reproduction number for the SARS-CoV-2 pandemic was difficult to determine due to two ‘saw-teeth’ in the data and the author was tempted to use only the troughs, nonetheless, didn’t. The basic reproduction number was calculated to be 3.16 (Table 1) and recent data suggests it has shot up, if it can be relied on. One also has to exercise caution in drawing conclusions from any comparison with South Africa. Sweden’s population-densities, their climate, their population’s way of life, etc. are very different to those in South Africa. One can nonetheless conclude that the *r*_0_ for the SARS-CoV-2 virus is high.

As much as a large proportion of the South African population live cheek by jowl, it is no credit to them that all lockdowns were characterised by a substantial lack of compliance. When one considers that similar lockdown regimes in countries like Australia and New Zealand produced the desired results, South Africans, to a certain extent need to blame themselves, as much as they do their circumstances, living conditions and government. The lockdowns served to ensure that the medical system was not overwhelmed, bought it valuable time to prepare and provided useful data. The lockdowns nonetheless failed significantly in meeting any objective to curtail the SARS-CoV-2 pandemic.

Despondence in the case of the SARS-CoV-2 virus can be tempered, to a limited extent, by contemplating the ‘flattening of the curve’ in the context of the Small-Epidemic Theorem. Severely reducing the infected fraction of the population just above the threshold could possibly alter the outcome. Knowing the exact, incorrect-data value, or perceived position, of this threshold could therefore be considered reasonably important and some statistician urgently needs to do some random testing to determine what the exact value of the multiplicative factor, *a*, really is.

## Data Availability

All data publically available, online, and listed in the bibliography.

